# Impact of social distancing regulations and epidemic risk perception on social contact and SARS-CoV-2 transmission potential in rural South Africa: analysis of repeated cross-sectional surveys

**DOI:** 10.1101/2020.12.01.20241877

**Authors:** Nicky McCreesh, Vuyiswa Dlamini, Anita Edwards, Stephen Olivier, Njabulo Dayi, Keabetswe Dikgale, Siyabonga Nxumalo, Jaco Dreyer, Kathy Baisley, Mark J. Siedner, Richard G. White, Kobus Herbst, Alison D. Grant, Guy Harling

## Abstract

**Background:** South Africa implemented rapid and strict physical distancing regulations to minimize SARS-CoV-2 epidemic spread. Evidence on the impact of such measures on interpersonal contact in rural and lower-income settings is limited.

**Methods:** We compared population-representative social contact surveys conducted in the same rural KwaZulu-Natal location once in 2019 and twice in mid-2020. Respondents reported characteristics of physical and conversational (‘close interaction’) contacts over 24 hours. We built age-mixing matrices and estimated the proportional change in the SARS-CoV-2 reproduction number (R_0_). Respondents also reported counts of others present at locations visited and transport used, from which we evaluated change in potential exposure to airborne infection due to shared indoor space (‘shared air’).

**Results:** Respondents in March-December 2019 (n=1704) reported a mean of 7.4 close interaction contacts and 196 shared air person-hours beyond their homes. Respondents in June-July 2020 (n=216), as the epidemic peaked locally, reported 4.1 close interaction contacts and 21 shared air person-hours outside their home, with significant declines in others’ homes and public spaces. Adults aged over 50 had fewer close contacts with others over 50, but little change in contact with 15-29 year olds, reflecting ongoing contact within multigenerational households. We estimate potential R_0_ fell by 42% (95% plausible range 14-59%) between 2019 and June-July 2020.

**Discussion:** Extra-household social contact fell substantially following imposition of Covid-19 distancing regulations in rural South Africa. Ongoing contact within intergenerational households highlighted the limitation of social distancing measures in protecting older adults.

**Funding:** Wellcome Trust, UKRI, DFID, European Union

## INTRODUCTION

The rapid spread of SARS-CoV-2 in 2020 has harmed populations both directly through Covid-19 morbidity and mortality, and indirectly via both less support for other health conditions ^1,2^ and economic impacts arising from government-imposed and self-directed reductions in social interaction.^3,4^ Local physical distancing regulations including mandatory ‘stay at home’ orders, restrictions on public gatherings, and banning of inter-household contact have been common during the pandemic.^5^ Such non-pharmaceutical interventions (NPI) were variably implemented and enforced in lower-income settings, notably in sub-Saharan Africa.^6^ Understanding the impact of both NPIs and personal decisions is vital to determining trade-offs between epidemic control and non-Covid wellbeing.

The impact of NPIs is likely to vary substantially across countries, reflecting differences in both demographic composition and social dynamics. There is particular concern that official movement limitations may have limited impact in settings where informal work is common and economic safety nets are limited.^7,8^ These concerns will be particularly important if the global pandemic follows the example of past infectious diseases and has its greatest impact on marginalized and previously disadvantaged populations.^9,10^

Quantitative data on relevant interpersonal interaction are central to such assessments, both directly for planning locally relevant evidence-based responses and for parameterisation of mathematical models of SARS-CoV-2 transmission and control policies. Movement data from sources linked to smartphones can indicate likely changes in contact patterns, they do not account for the detailed, non-random social interaction that often typifies human behaviour,^11^ and in settings with low smartphone penetration, reliance on such data can lead to biased results.

Detailed quantitative social contact surveys have been conducted during the Covid-19 pandemic, primarily in higher-income settings. These include online surveys using de novo convenience recruitment in Europe,^12,13^ and existing online panels in Europe ^14,15^ and the United States.^16^ Telephonic surveys have been conducted using random digit dialling in China,^17^ and existing cohorts in Kenya.^18^

Maximizing the benefit of these Covid-19 social contact studies requires careful study design. First, a clear sampling frame rather than a convenience sample allows stronger inference to a source population. Second, having comparable pre-pandemic data allows a clear measure of change to be assessed – there is danger of recall bias if questions are asked retrospectively about pre-pandemic days, and of secular change prior to Covid-19 if using previously collected data from too long ago. Third, longitudinal data within the epidemic’s progress allows judgement of the effects of changing policy and compliance willingness. Fourth, given evidence for aerosolized SARS-CoV-2 transmission,^19^ adding information on contact-time occurring in indoor congregate settings and transport can broaden our understanding of risk.

South Africa implemented an early, stringent national lockdown in March 2020, which may have initially delayed the national epidemic;^20^ however regulations were relaxed from May onwards and case numbers increased rapidly, peaking in July before falling back. In this paper, we compare data from two studies conducted using comparable study instruments in 2019 and 2020, both using samples drawn from the same census sampling frame in rural South Africa. The 2020 data include two rounds of data collection, covering the first wave of Covid-19 in the local area. We use these data to estimate the reduction in potential reproduction number of SARS-CoV-2 between surveys, and to determine where contact beyond the home continued during lockdown periods.

## METHODS

We used data from two surveys conducted in the southern section of the Africa Health Research Institute (AHRI) demographic surveillance area in 2019 and 2020. AHRI maintains an active thrice-yearly census of all households ∼21,000 households in this area of ∼850km^2^ in rural uMkhanyakude district, KwaZulu-Natal province, including one small town.^21^ uMkhanyakude ranks among the most deprived districts nationally in terms of health and socioeconomic status.

The 2019 data were collected as part of Umoya omuhle (UO), a programme exploring novel approaches to prevention of drug-resistant *Mtb* transmission in health facilities.^22^ UO sampled 3093 census adults (aged 18 and above) residing within the census surveillance area and the catchment area of two primary care centres (one in town, one rural). Sampling was random, stratified by residential area (∼350 households per area) and with probability proportional to the number of eligible people in each area, based on the most recent census conducted prior to area entry. Data collection was conducted March to December 2019 at respondents’ homes.

The 2020 data were collected as part of a longitudinal Covid-19 surveillance project.^23^ The Covid Social Contact (CSC) sub-study used an age/sex stratified sample of one person aged 15 and above from each of 400 census households. Inclusion criteria included participation in Vuk’uzazi, a recent population-wide chronic health screening study,^24^ allowing intentional oversampling of individuals with locally prevalent health conditions (tuberculosis, hypertension, diabetes, obesity, COPD or asthma). Contact was made telephonically based on previously provided numbers. We analysed two rounds of data collected between 3 June and 16 July (2020 R1), and 16 July and 17 August (2020 R2), as Covid-19 peaked in KwaZulu-Natal (Figure 1).

**Figure 1:**
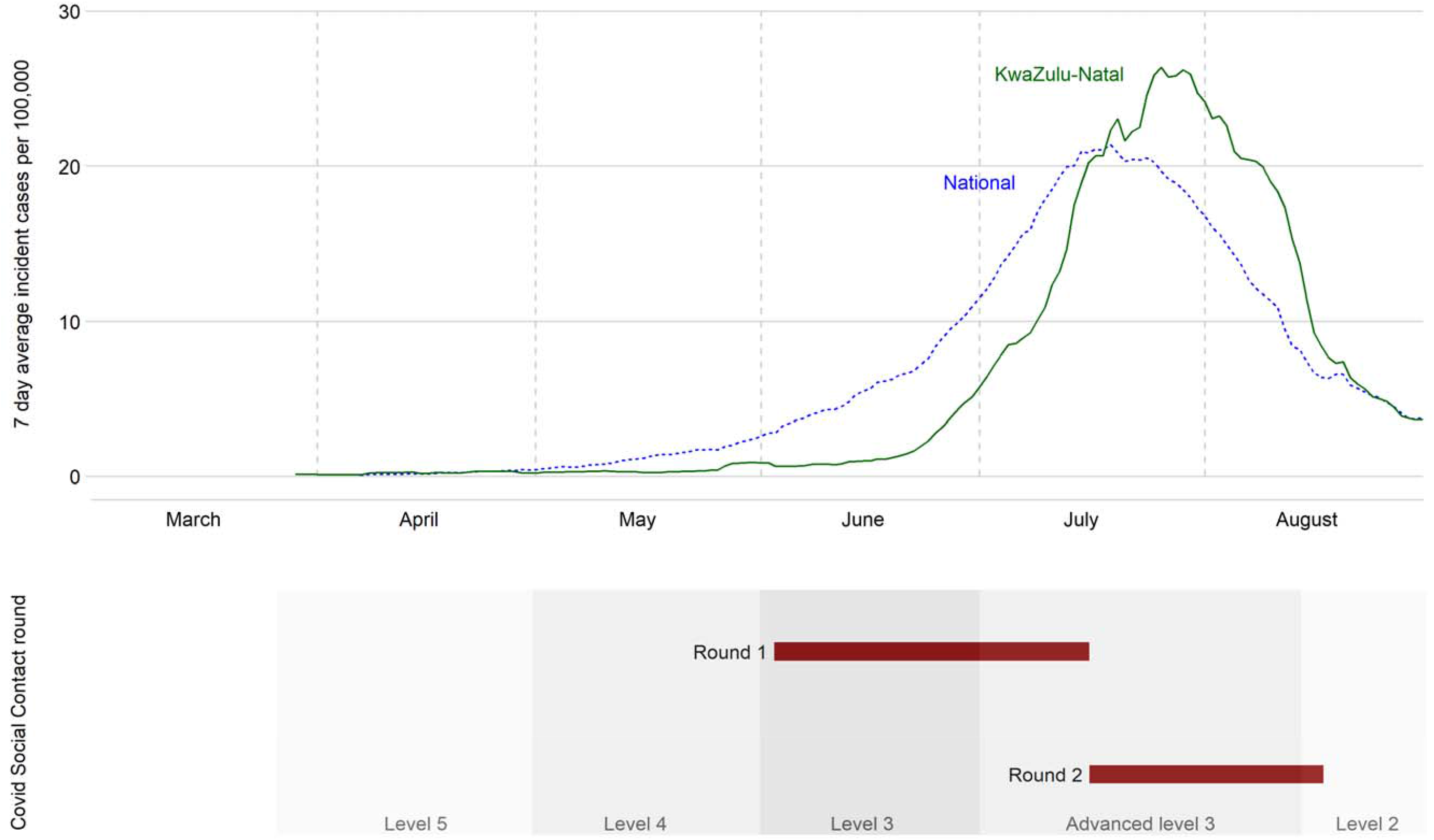
South African smoothed Covid-19 case incidence rate, government lockdown and 2020 survey dates in 2020. Levels refer to national non-pharmaceutical interventions (“lockdown”) with strictness declining from level 5 over time. Incidence rates are 7-day running mean values computed from data collated at https://mediahack.co.za/datastories/coronavirus/dashboard/.

Both studies used structured electronic REDCap interviews (Supplementary Material 1). After confirming socio-demographic characteristics, respondents were asked to report on three forms of in-person social interaction. First, they were asked to list all indoor locations visited over a 24-hour period. UO asked about a randomly selected day in the past week; CSC asked about the day prior to interview, limiting data largely to Sunday to Thursday. Follow up questions covered the type of location, length of time spent there and the number of people present. Second, respondents were asked who they had directly interacted with over the 24-hour period – involving either physical contact or a minimum-three-word conversation. Respondents were then for each ‘close interaction contact’ asked to report their ages and sexes and the duration of time spent together; UO asked these follow-up questions about a random 10 contacts, or all contacts if 10 or fewer were reported while CSC asked about all contacts. Third, respondents were asked about any transport used over the past day, and then how long any trips took and many people shared the transport with them. CSC only asked buildings and transport questions to a random half of sampled individuals per round.

### Statistical analyses

We grouped respondents and their close interaction contacts by age into four categories (15-29, 30-49, 50-64 and 65+) and described respondent characteristics for each survey round. All subsequent analyses weighted the data for sampling and non-response to match the census population and to make methods comparable across surveys (further details in Supplementary Material 2). We first calculated the mean number of close interaction contacts per person per day by respondent characteristics (sex, age, household size) and by whether the contact was a household member. We then built social contact age mixing matrices for each round, adjusting our raw results to ensure that the matrices were symmetric using the census population age structure. From these values we calculated the change between 2019 and each 2020 round.

In the absence of a robust estimate of the basic reproduction number (R_0_) for SARS-CoV-2 in rural South Africa without social distancing (i.e., with 2019 social contact patterns), we estimated the relative reduction in R_0_ between rounds, assuming the per-contact transmission probability remained constant. (Actual reductions will have been greater to the extent that face coverage increased in 2020 but we cannot precisely estimate either the degree to which this occurred, especially within households, or the degree of protection conferred.) For our estimated reduction we used the next generation matrix of the age-contact matrix, defining R_0_ as the dominant eigenvalue. To assess uncertainty, we generated mean and 95% plausible intervals using 10,000 independent bootstrapped samples of each survey, and calculated the relative reduction in each pair of samples. The bootstrapped samples were generated by re-sampling respondents with replacement within age categories, and re-sampling contacts with replacement from the set of all contacts of the respondent.^25^

Finally, we calculated the change in potential exposure to airborne infection using location and transport data by calculating for each respondent per day: the proportion who visited any location/transport type; mean hours spent in the location if visited; mean people present per visit; mean ‘shared air person-hours’ if visited; and finally mean shared air person-hours across all respondents. For this analysis we merged the two CSC rounds since each respondent provided one datapoint. We tested for significant differences in each measure using logistic or linear bivariate regression including indicator variables for study round.

We conducted several sensitivity analyses: i) including only data on Sundays to Thursdays, giving each day equal weight, to compare only same-day data across the two surveys; ii) excluding respondents with any missing close interaction contact age data; iii) excluding close interaction contacts of less than 15 minutes duration, as the probability of transmission is lower for shorter contact durations; iv) incorporating children into our analysis, using UO and CSC data on adult-reported contact with children, and past South African data about child-child contacts; (v) including only individuals aged ≥18, to make the two datasets comparable on age range; (vi) limiting UO data to the period June-August 2019 to ensure seasonal comparability; (vii) weighted the 2019 data to the full census population based on urbanicity (further details in Supplementary Material 2).

Ethical approval for UO was granted by the Biomedical Research Ethics Committee (REC) of the University of KwaZulu-Natal (UKZN) (BE662/17) and the London School of Hygiene & Tropical Medicine (14640); ethical approval for CSC was granted by UKZN BREC (BE290/16) and University College London REC (15231/013). Informed consent for participation was recorded in writing for UO and telephonically for CSC.

## RESULTS

Of the 3093 people sampled for UO, 1723 (56%) were successfully contacted, 299 (10%) were dead or reported to have out-migrated, and 1071 (35%) could not be contacted. Of those successfully contacted, 1704 (99%) completed an interview. Of the 400 individuals sampled for CSC, 27 (7%) were dead or had out-migrated, and 102 (26%) could not be contacted. Of those successfully contacted, 216 (80%) completed an interview. At R2 follow-up, 202 of the 216 (94%) completed a second interview and eight previously uncontactable individuals were reached for a first interview. The raw age-sex structures of UO and CSC differed from one-another by design (Table 1).

**Table 1:** Descriptive statistics of respondents to the Umoya omuhle (2019) and Covid Social Contacts (2020) studies.

|  | 2019 |  | 2020 R1: June-July |  |  |  | 2020 R2: July-August |  |  |  |  |
| --- | --- | --- | --- | --- | --- | --- | --- | --- | --- | --- | --- |
|  | All |  | Close contact |  | Buildings & transport |  | Close contact |  | Buildings & transport |  | Census population |
| Age |  |  |  |  |  |  |  |  |  |  |  |
| 15-29 | 613 | 36% | 36 | 17% | 15 | 14% | 32 | 15% | 17 | 17% | 42% |
| 30-49 | 535 | 31% | 50 | 23% | 28 | 26% | 47 | 22% | 23 | 23% | 33% |
| 50-64 | 342 | 20% | 64 | 30% | 35 | 33% | 65 | 31% | 28 | 28% | 16% |
| 65+ | 214 | 13% | 66 | 31% | 29 | 27% | 66 | 31% | 33 | 33% | 10% |
| Sex |  |  |  |  |  |  |  |  |  |  |  |
| Male | 751 | 44% | 101 | 47% | 49 | 46% | 97 | 46% | 48 | 48% | 42% |
| Female | 953 | 56% | 115 | 53% | 58 | 54% | 113 | 54% | 53 | 52% | 58% |
| Household size |  |  |  |  |  |  |  |  |  |  |  |
| 1-3 | 293 | 17% | 61 | 28% | 33 | 31% | 62 | 30% | 27 | 27% | 19% |
| 4-6 | 426 | 25% | 60 | 28% | 28 | 26% | 60 | 29% | 29 | 29% | 34% |
| 7-9 | 429 | 25% | 54 | 25% | 29 | 27% | 48 | 23% | 22 | 22% | 25% |
| 10+ | 556 | 33% | 41 | 19% | 17 | 16% | 40 | 19% | 23 | 23% | 22% |
| Residence |  |  |  |  |  |  |  |  |  |  |  |
| Urban/peri-urban | 837 | 49% | 85 | 39% | 37 | 35% | 84 | 40% | 41 | 41% | 32% |
| Rural | 867 | 51% | 131 | 61% | 70 | 65% | 125 | 60% | 59 | 58% | 59% |
| Unknown | 0 |  | 0 |  | 0 |  | 1 | 0.5% | 1 | 1.0% | 0.4% |
| Day reported |  |  |  |  |  |  |  |  |  |  |  |
| Monday | 239 | 14% | 49 | 23% | 22 | 21% | 25 | 12% | 15 | 15% |  |
| Tuesday | 242 | 14% | 52 | 24% | 24 | 22% | 26 | 12% | 12 | 12% |  |
| Wednesday | 239 | 14% | 37 | 17% | 20 | 19% | 73 | 35% | 35 | 35% |  |
| Thursday | 251 | 15% | 40 | 19% | 24 | 22% | 70 | 33% | 33 | 33% |  |
| Friday | 261 | 15% | 0 |  | 0 |  | 1 | 0% | 1 | 1% |  |
| Saturday | 245 | 14% | 0 |  | 0 |  | 0 |  | 0 |  |  |
| Sunday | 227 | 13% | 38 | 18% | 17 | 16% | 15 | 7% | 5 | 5% |  |
| Total | 1704 |  | 216 |  | 107 |  | 210 |  | 101 |  | 36 311 |

Values are counts and percentages. Census population refers to proportions of residents aged 15+ in the area.

The mean number of close interaction contacts varied little by respondent age, sex or household size within rounds, although numbers were lower in the highest age group, and were positively associated with household size (Table 2). Respondents reported a mean of 7.4 close interaction contacts in 2019 (95%CI: 7.1-7.7), 4.1 (95%CI: 3.5-4.6) in 2020 R1 and 4.3 (95%CI: 3.8-4.8) in 2020 R2. Contact reductions were larger for non-household than household member contacts in both absolute and relative terms. Non-household contacts fell from 2.8 (95%CI 2.6-3.1) in 2019 to 0.7 (95%CI 0.4-1.1) in 2020 R1 and 0.5 (95% CI 0.3-0.7) in 2020 R2. Household contacts fell from 4.6 (95% CI 4.5-4.8) in 2019 to 3.4 (95% CI 2.9-3.8) in 2020 R1 and 3.8 (95% CI 3.3-4.3) in 2020 R2.

**Table 2:** Mean close contact numbers by respondent characteristics.

|  | 2019 |  | 2020 R1 |  | p-value | 2020 R2 |  | p-value |
| --- | --- | --- | --- | --- | --- | --- | --- | --- |
| **All contacts** | 7.4 | [7.1 - 7.7] | 4.2 | [3.5 - 5.0] | <0.001 | 4.3 | [3.3 - 5.8] | <0.001 |
| Age |  |  |  |  |  |  |  |  |
| 15-29 | 8.1 | [7.6 - 8.6] | 4.7 | [3.3 - 6.8] | <0.001 | 4.6 | [3.5 - 5.9] | <0.001 |
| 30-49 | 7.0 | [6.7 - 7.4] | 3.6 | [2.9 - 4.4] | <0.001 | 4.0 | [3.2 - 5.1] | <0.001 |
| 50-64 | 7.2 | [6.5 - 8.0] | 4.8 | [3.7 - 6.2] | 0.001 | 4.8 | [3.7 - 6.2] | 0.001 |
| 65+ | 6.1 | [5.5 - 6.7] | 3.3 | [2.7 - 4.0] | <0.001 | 3.4 | [2.7 - 4.2] | <0.001 |
| Sex |  |  |  |  |  |  |  |  |
| Male | 7.1 | [6.7 - 7.5] | 4.6 | [3.3 - 6.3] | 0.001 | 3.4 | [2.6 - 4.5] | <0.001 |
| Female | 7.6 | [7.3 - 8.0] | 4.0 | [3.3 - 4.7] | <0.001 | 4.9 | [4.2 - 5.6] | <0.001 |
| Household size |  |  |  |  |  |  |  |  |
| 1-3 | 6.1 | [5.4 - 6.9] | 2.6 | [2.0 - 3.3] | <0.001 | 2.2 | [1.4 - 3.4] | <0.001 |
| 4-6 | 6.9 | [6.4 - 7.4] | 4.0 | [3.4 - 4.8] | <0.001 | 4.0 | [3.3 - 4.8] | <0.001 |
| 7-9 | 7.6 | [7.1 - 8.2] | 4.2 | [3.4 - 5.1] | <0.001 | 4.7 | [3.9 - 5.7] | <0.001 |
| 10+ | 8.3 | [7.9 - 8.8] | 5.8 | [3.6 - 9.2] | 0.054 | 6.0 | [4.6 - 7.7] | 0.002 |
| Residence |  |  |  |  |  |  |  |  |
| Urban/peri-urban | 7.1 | [6.7 - 7.5] | 4.5 | [3.6 - 5.7] | <0.001 | 4.4 | [3.3 - 5.8] | <0.001 |
| Rural | 7.7 | [7.3 - 8.1] | 4.0 | [3.1 - 5.2] | <0.001 | 4.4 | [3.8 - 5.0] | <0.001 |
| **Household members** | 4.6 | [4.5 - 4.8] | 3.4 | [3.0 - 3.9] | <0.001 | 3.8 | [3.3 - 4.4] | 0.01 |
| Age |  |  |  |  |  |  |  |  |
| 15-29 | 4.8 | [4.6 - 5.1] | 3.6 | [2.7 - 4.8] | 0.018 | 4.1 | [3.0 - 5.4] | 0.19 |
| 30-49 | 4.4 | [4.2 - 4.7] | 2.9 | [2.3 - 3.6] | <0.001 | 3.6 | [2.7 - 4.6] | 0.066 |
| 50-64 | 4.5 | [4.2 - 4.9] | 4.2 | [3.3 - 5.5] | 0.58 | 4.0 | [3.2 - 5.1] | 0.032 |
| 65+ | 4.6 | [4.1 - 5.1] | 3.0 | [2.5 - 3.7] | <0.001 | 3.1 | [2.5 - 3.9] | 0.001 |
| Sex |  |  |  |  |  |  |  |  |
| Male | 3.9 | [3.7 - 4.1] | 3.3 | [2.7 - 4.1] | 0.090 | 2.7 | [2.0 - 3.6] | 0.002 |
| Female | 5.1 | [4.9 - 5.4] | 3.5 | [2.9 - 4.2] | <0.001 | 4.5 | [3.9 - 5.2] | 0.073 |
| Household size |  |  |  |  |  |  |  |  |
| 1-3 | 3.2 | [2.9 - 3.5] | 1.6 | [1.2 - 2.2] | <0.001 | 1.5 | [1.0 - 2.4] | <0.001 |
| 4-6 | 3.9 | [3.7 - 4.2] | 3.4 | [2.9 - 3.9] | 0.025 | 3.3 | [2.7 - 4.2] | 0.11 |
| 7-9 | 4.8 | [4.6 - 5.1] | 3.8 | [3.1 - 4.6] | 0.012 | 4.5 | [3.7 - 5.5] | 0.50 |
| 10+ | 5.7 | [5.4 - 6.1] | 4.5 | [3.1 - 6.6] | 0.15 | 5.3 | [4.1 - 6.8] | 0.52 |
| Residence |  |  |  |  |  |  |  |  |
| Urban/peri-urban | 4.3 | [4.1 - 4.6] | 3.8 | [2.9 - 4.9] | 0.298 | 3.7 | [2.7 - 5.0] | 0.26 |
| Rural | 5.0 | [4.8 - 5.3] | 3.3 | [2.8 - 3.9] | <0.001 | 4.0 | [3.4 - 4.7] | 0.00 |
| **Non-household members** | 2.8 | [2.6 - 3.1] | 0.78 | [0.44 - 1.4] | <0.001 | 0.52 | [0.52 - 0.32] | <0.001 |
| Age |  |  |  |  |  |  |  |  |
| 15-29 | 3.3 | [2.9 - 3.7] | 1.2 | [0.42 - 3.2] | 0.00 | 0.52 | [0.52 - 0.19] | <0.001 |
| 30-49 | 2.7 | [2.4 - 3.0] | 0.69 | [0.40 - 1.2] | <0.001 | 0.48 | [0.48 - 0.20] | <0.001 |
| 50-64 | 2.7 | [2.0 - 3.5] | 0.53 | [0.24 - 1.1] | <0.001 | 0.76 | [0.76 - 0.39] | <0.001 |
| 65+ | 1.6 | [1.2 - 2.1] | 0.28 | [0.14 - 0.54] | <0.001 | 0.26 | [0.26 - 0.10] | <0.001 |
| Sex |  |  |  |  |  |  |  |  |
| Male | 3.3 | [2.9 - 3.7] | 1.3 | [0.53 - 3.1] | 0.00 | 0.79 | [0.79 - 0.41] | <0.001 |
| Female | 2.5 | [2.2 - 2.9] | 0.48 | [0.31 - 0.75] | <0.001 | 0.36 | [0.36 - 0.18] | <0.001 |
| Household size |  |  |  |  |  |  |  |  |
| 1-3 | 3.0 | [2.4 - 3.8] | 1.0 | [0.59 - 1.6] | <0.001 | 0.62 | [0.62 - 0.25] | <0.001 |
| 4-6 | 3.0 | [2.5 - 3.5] | 0.66 | [0.38 - 1.2] | <0.001 | 0.68 | [0.68 - 0.32] | <0.001 |
| 7-9 | 2.8 | [2.4 - 3.4] | 0.37 | [0.16 - 0.85] | <0.001 | 0.20 | [0.20 - 0.06] | <0.001 |
| 10+ | 2.6 | [2.3 - 3.0] | 1.3 | [0.32 - 5.1] | 0.13 | 0.67 | [0.67 - 0.23] | <0.001 |
| Residence |  |  |  |  |  |  |  |  |
| Urban/peri-urban | 2.8 | [2.5 - 3.1] | 0.73 | [0.45 - 1.2] | <0.001 | 0.71 | [0.71 - 0.38] | <0.001 |
| Rural | 2.7 | [2.4 - 3.0] | 0.70 | [0.24 - 2.1] | <0.001 | 0.39 | [0.39 - 0.16] | <0.001 |

p-values are for comparisons within rows vs. 2019 data.

In 2019 weighted age-mixing matrices, older individuals had fewer close interaction contacts than younger ones, and contact rates for each age group under 65 were highest within the same age group. By June/July 2020 (2020 R1), contact rates were lower for all age combinations, in most cases statistically significantly. The drop was greatest for contact between those aged 65+, and smallest for contact between those aged 15-29 and 50-64. The drops in contacts adults reported having with children were lower than for between adults, and in most adult age groups the data were consistent with no change in contact rates between adults and children (Supplementary Material 3, panel 4). Contact patterns by age changed little between 2020 R1 and R2. The estimated reduction in R_0_ between 2019 and 2020 R1 was 45% (95% plausible range 14-59%), and between 2019 and 2020 R2 was 45% (95% plausible range 24-61%) (Figure 2a-c).

**Figure 2:**
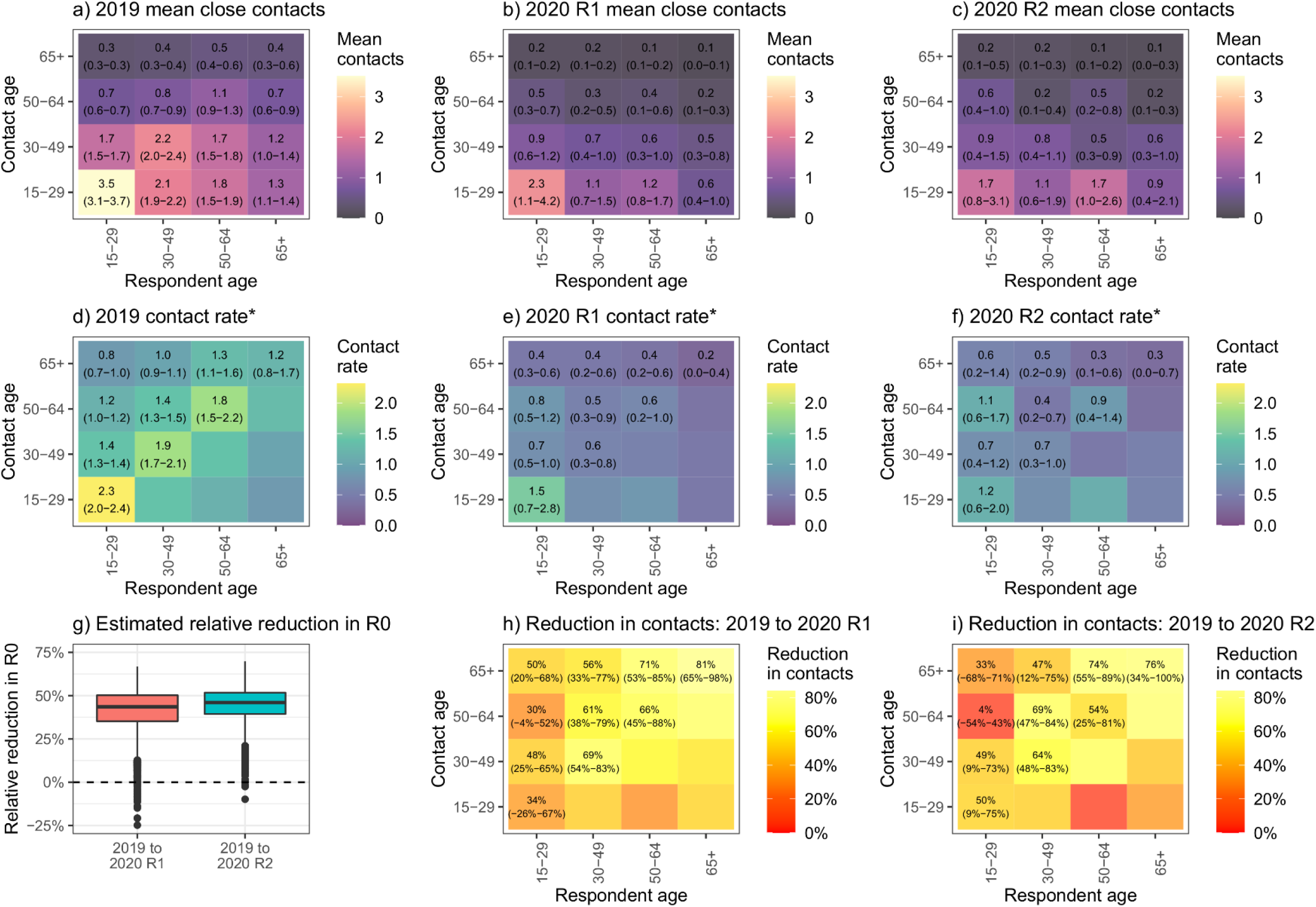
Age-stratified mean (95% plausible range) number of close contacts from survey respondents. Graphs a-c show the mean number of contacts respondents in each age group reported, by contact age group. Graphs d-f show the mean rate of contact between respondents in each age group and each other person in the target population, by contact age group. *numbers shown are rates x 10^5^.

Sharing of indoor space more generally also fell between 2019 and mid-2020: mean shared air person-time beyond respondents’ own homes (including transport) fell from 196 to 21 hours (Table 3). Mean time spent at one’s own home rose by almost three hours per day, from an average of 19 to 22 hours, although there was no significant change in person-contact hours. All other location types except public transport saw substantially reduced overall person-contact hours, largely due to fewer people present in the location, rather than changes in the time spent per visit. The proportion of people reporting clinic visits almost doubled, from 2.3% to 4.3%, although the change was not significant (p=0.20). The proportion of respondents who reported visiting other people’s households and ‘other’ locations fell, from 27% to 6%, and 32% to 24%, respectively.

**Table 3:**
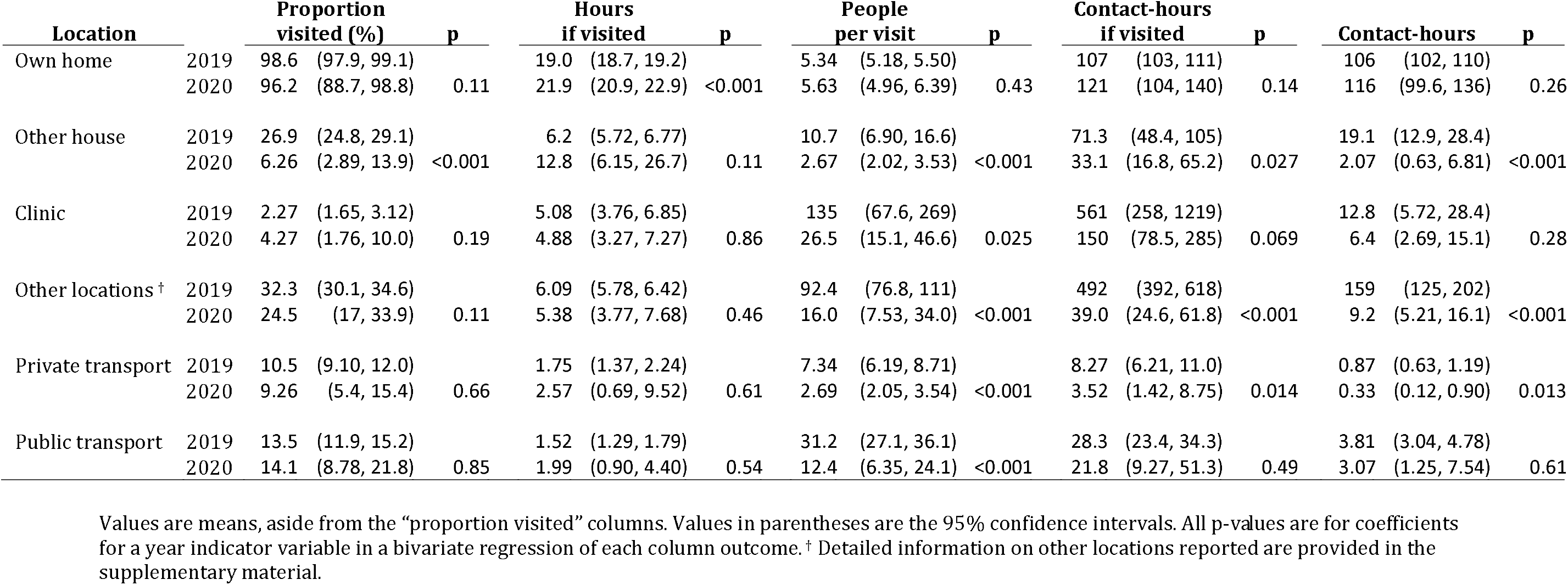
Location and transport patterns of respondents in 2019 and 2020.

Our sensitivity analyses did not substantively change our conclusions (Supplementary Material 3). Estimated reductions in R_0_ between 2019 and 2020 R1 and 2020 R2 ranged from 40-50% and 38-48% respectively in the sensitivity analyses, compared to 42% and 45% respectively in the primary analysis. Excluding Fridays/Saturdays, contacts under 15 minutes, respondents with any missing contact age data or those aged 15-17 had no qualitative effect on age-mixing patterns. Adding children to the analysis highlighted the large number of contacts adults had with children pre-Covid, but did not affect the estimated reduction in R_0_ between 2019 and 2020. However, it was notable that adults’ close personal contact with children fell by less than almost any other age combination, looking similar to the pattern between 15-29 and 50-64 year olds.

## DISCUSSION

We compared rates of close interaction contact and time spent with others indoors or on transport from surveys conducted in the same rural South African setting, both pre-Covid-19 in 2019 and during the first wave of cases in mid-2020. We found substantial declines in close contact numbers and in time spent at most indoor locations other than respondents’ own homes by 2020, suggesting that the combination of government NPIs and the ongoing epidemic substantially affected behaviour. Under the assumptions that close interaction contacts are the most important for infection transmission, and qualitatively the same in both years, we estimate a 42-45% reduction in the likely basic reproduction number between the two surveys. Given the substantial level of use of face coverings outside the home required and observed in South Africa,^26^ this reduction is likely to be an underestimate.

While it is difficult to disentangle the effects of NPIs and the state of the local epidemic, the fact that much of the decline between 2019 and 2020 was present by the first interview round – the majority of which occurred prior to the local mass arrival of Covid-19 – suggests that respondents in this area were complying with government mobility NPIs. By June and July these had been relaxed somewhat from the initial strict stay-at-home requirements, but entertainment and alcohol availability remained highly limited. The continuation of limited close interaction contact, even as NPIs were relaxed into August and beyond, highlights that the changes made in response to NPIs were maintained subsequently; it is hard to tell whether this was due to increased concern due to widespread local transmission or slow adjustment to changing policies.

Declines in close interaction contact were not homogenous. The age group with the greatest decreases in contact were those aged over 65, particularly for contact with others aged 50 and older, with declines of around three-quarters compared to pre-Covid. In contrast, the lowest declines were seen between those aged 15-29 and older adults – particularly those aged 30-49 years, i.e. one generation older. This pattern reflects the notably larger decline in non-household member contacts compared to household members in combination with the common presence of multiple adult generations within each household. This finding is similar that seen in other contact studies of rural Africans ^27^, and highlights the likely difficulty of protecting those most vulnerable to Covid-19 in settings such as this. A fuller understanding of the implications of these ongoing within-household intergenerational contacts will require focused qualitative work to determine whether young adults living in multigenerational households are able to maintain some social distance within houses, and how to potentially target messages to this population.

To consider the implications of behaviour change for an infection with aerosol transmission potential, we also measured time spent in indoor locations, and numbers of people present. Unsurprisingly, we found that time spent at home rose, while shared person-time spent in other homes dropped by 89% and at ‘other locations’ (largely school, work and shops) by 94%. This reduced potential for exposure in both public and private suggests that people are following rules both when they can and cannot be seen; these data are also consistent with other evidence from this area of reduced mobility in July and August compared even to earlier in 2020 ^26^. Our respondents reported similar time spent attending health clinics in the two years, but reported that fewer others were present when they attended in 2020, perhaps reflecting improved social distancing policies within clinics. Overall, our indoor location data suggested nuanced decision-making by respondents during Covid-19, reducing less vital trips but maintaining necessary ones.

Even prior to Covid-19, the numbers of close interaction contacts reported in this area was substantially lower than that seen elsewhere in Africa.^18,27-30^ Low numbers of contacts may reflect the very large proportion of our respondents’ days spent at home – a mean of 19 hours in 2019 and almost 22 hours in 2020. This lack of mobility reflects very high local unemployment – under 25% of resident working-aged adults reported employment during the late-2019 demographic census. As a result, while household contact numbers are comparable to other African studies, contacts beyond the household appear to be substantially lower. In the context of a South African first wave of the Covid-19 epidemic that saw much smaller outbreaks in rural than urban areas, this lower baseline suggests a low rural R_0_ even before individuals increased their social distance.

### Strengths and limitations

There are limitations to this study. As with any observational study of human behaviour, care must be taken in generalising to other populations. While the pre-Covid-19 contact patterns we show here are consistent with those elsewhere in rural Africa,^27^ as are the changes seen with the arrival of Covid-19,^18^ it is important to consider whether close contact patterns in rural lower-income settings may have different implications for disease prevention than patterns seen elsewhere.

In contrast to social contact surveys that used prospective diaries to capture information, we relied on recent recall – this may have led to some misreporting, but the delay was in all cases less than one week, limiting this concern. Our data were also self-reported rather than, for example, based on proximity detectors or mobility tracking. Self-report can lead to misreporting, although this effect is unlikely to have affected measures of change since we used the same approach for both years. Self-report also has the benefit of providing richer data on the nature of each interaction. Conversely, we kept our questionnaires brief to minimize respondent fatigue, and as a result we do not have certain details about each contact, including whether a facemask was used during each interaction.

There are substantial strengths to this work. We were able to compare two studies with respondents drawn from the same well-defined sampling frame asking very similar questions about behaviour both shortly before and after the arrival of Covid-19 in the study area. The availability of longitudinal response data during the Covid-19 outbreak allowed us to observe behaviour changed as government NPIs and epidemic situations changed; although while patterns of behaviour over time can suggest effects, we cannot prove causality, something important if using our findings to design preventative interventions.

## Conclusion

In comparable surveys about social contacts conducted in the same rural South African location in 2019 and mid-2020, we find substantial declines in close physical and conversational contacts, and also in beyond-household sharing of indoor space. These findings suggest that the strict government NPIs implemented to mitigate the Covid-19 epidemic, in combination with the arrival of the epidemic itself in the local area, led to highly protective behaviours. It will be important to triangulate these findings with other information on the wider impact of such behaviour.

## Supporting information

STROBE checklist

## Data Availability

Individual respondent data that underlie the results reported in this article, after de-identification, will be made available immediately following publication indefinitely at https://data.ahri.org/ for anyone providing a methodologically sound proposal to conduct analysis.

## Acknowledgements

We acknowledge the generosity of all participants. We gratefully acknowledge the efforts of all the study research assistants: in 2019 Nkosingiphile Buthelezi, Zilethile Khumalo, Sifundesihle Malembe, Zodwa Mkwanazi, Sanele Mthiyane; in 2020 Gugu Buthelezi, Ngenzeni Buthelezi, Duduzile Mkhwanazi, Siyabonga Mnyango, Nondumiso Mpanza, Mxolisi Nhlenyama. We thank Indira Govender for helpful suggestions on an earlier draft.

## Funding Statements

Data collection in the Umoya omuhle programme is funded in part by the UK Economic and Social Research Council (ES/P008011/1), one of seven research councils underpinning the Antimicrobial Resistance Cross Council Initiative. It is also supported by the UK Medical Research Council (MRC) and the UK Department for International Development (DFID) under the MRC/DFID Concordat agreement that is also part of the EDCTP2 programme supported by the European Union (MR/P002404/1). The Covid Social Contacts study was funded by internal seed funding from the Africa Health Research Institute (AHRI), which is core funded by the Wellcome Trust (201433/Z/16/Z).

GH is supported by a fellowship from the Royal Society and the Wellcome Trust (210479/Z/18/Z). NM, RGW and ADG are supported by the UK ESRC (ES/P008011/1); NM and RGW are supported by the Wellcome Trust (218261/Z/19/Z). RGW is additionally funded NIH (R01-AI147321), EDTCP (RIA208D-2505B), UK MRC (CCF17-7779 via SET Bloomsbury), BMGF (OPP1084276, OPP1135288 & INV-001754), and the WHO (2020/985800-0).

## Declaration of Interests

The authors have no conflicts of interest to declare.

## Authors’ contributions

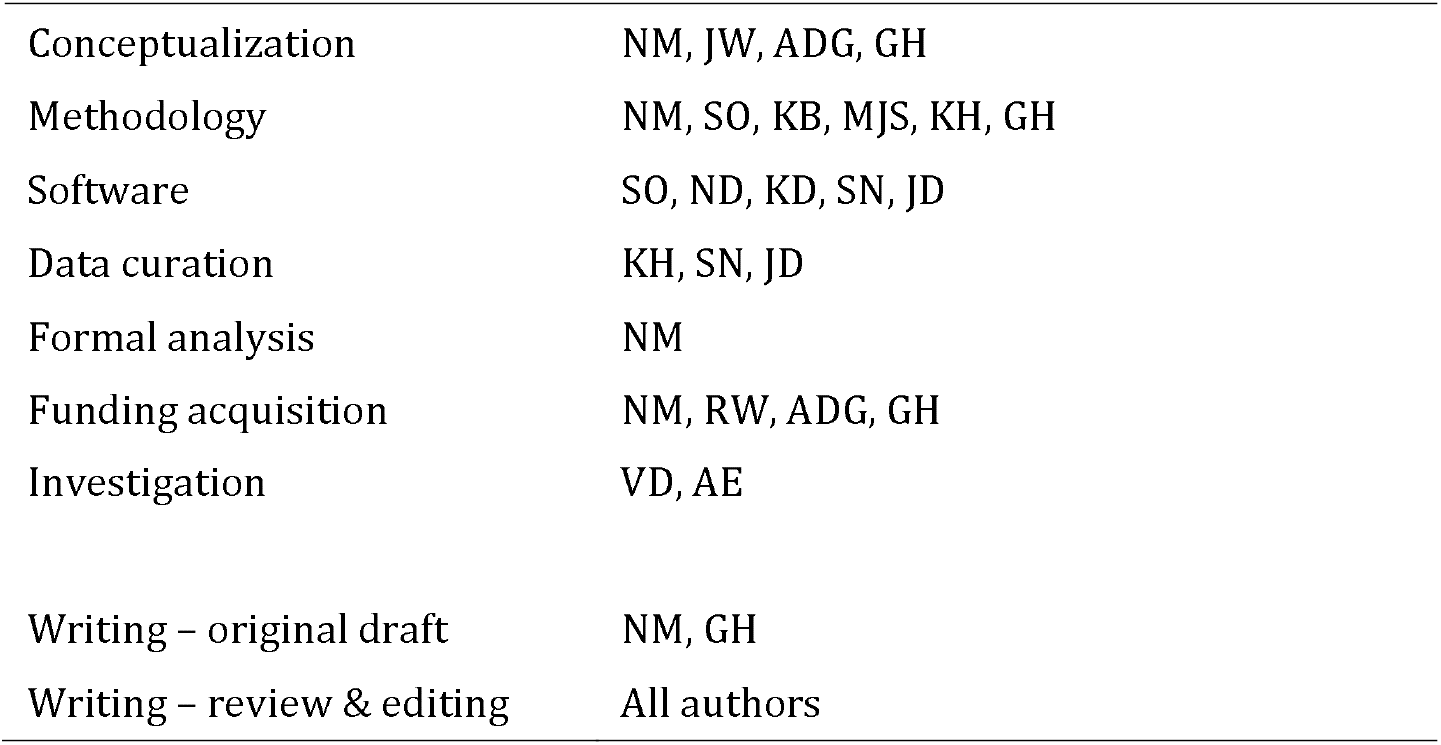

All authors approved the final version and agree to be accountable for the work.

## Supplementary Material

Title: Impact of social distancing regulations and epidemic risk perception on social contact and SARS-CoV-2 transmission potential in rural South Africa: analysis of repeated cross-sectional surveys

### Supplementary Material 1. Study instruments

|  | Physical locations |  |
| --- | --- | --- |
|  | Make a list of all the indoor places you used on [day] between 12am and 12am*. The interviewer will make a list of all of the places on a piece of paper. This will only be used to help you remember the places. Do not include locations that you did not go inside (e.g. shops where you were served through a hatch). Include tents that were enclosed, but not tents that only had a roof. This should include your own home if you spent time there. If you visited a location more than once, then list it more than once, and answer the subsequent questions for each time you visited. For workplaces, put the type of workplace (e.g. shop). |  |
| ph01 | How many places are on your list of buildings visited? | Integer |
|  | [Loop over list from PH01 for PH02-PH07] |  |
| ph02 | What type of location was it? | 0 "Own home" 1 "Other house on plot" 2 "House off plot" 3 "Church" 4 "Clinic" 5 "Community building" 6 "Creche" 7 "Gym" 8 "Library" 9 "Mall / Shops" 10 "in Bar/ Nightclub" 11 "Spaza" 12 "School" 13 "Salon/Barber" 14 "In a tent" 15 "Counselling Centre" 16 "Guest House" 17 "Office" 18 "Workshop" 19 "Restaurant" 96 "Other" 98 "Prefer not to say" |
| ph03<sup>†</sup> | Which isigodi (local area) was it in? | [List of izigodi (local areas)] |
| ph04<sup>†</sup> | Is this place your workplace? | 0 "No" 1 "Yes" 99 "Don't know" 98 "Prefer not to say" |
| ph05 | What time did you arrive? | Time |
| ph06 | How long did you spend there? | time |
| ph07 | How many people (adults and children) were there, halfway through the time you were there? | integer |
| ph08 | How many of those people were children aged < 15? | Integer |
| ph09 | On how many days did you visit this location in the past week? | integer |
|  | Make a list of all the transport you used on [day] between 12am and 12am*. If you used a method of transport more than once (for instance a trip on two different taxis), then list it more than once, and answer the subsequent questions for each time you used it. |  |
| ph10 | How many trips are on your list of trips made? | integer |
|  | [Loop over list from PH8 for PH9-13] |  |
| ph11 | What type of transport was it? | 0 "Taxi" 1 "Bakkie" 2 "Private car" 3 "Bus" 4 "Motorbike" 5 "Bicycle" 96 "Other" 98 "Prefer not to answer" |
| ph12<sup>†</sup> | Where did it start? | [List of izigodi (local areas)] |
| ph13<sup>†</sup> | Where did it end? | [List of izigodi (local areas)] |
| ph14 | What time did it start? | time |
| ph15 | How long did the journey take? (Don't worry if approximate) | time |
| ph16 | How many people (adults and children) were on the vehicle at the start of your trip? | integer |
| ph17 | How many of those people were children aged < 15? | integer |
| ph18<sup>†</sup> | Is this the transport you typically use on [day]? |  |
| ph19<sup>†</sup> | [If PH18 == 0] Do you usually make no trips using transport on [day]s? | 0 "No" 1 "Yes" 99 "Don't know" 98 "Prefer not to say" |
| **Social contacts 2019** |  |  |
| sc01 | Make a list of all the people you spoke to or touched on [day]. Make a note of which are in your household. Include face-to-face meetings only (phone calls don't count). By [day], we mean between 12am and 12am.<br>The interviewer will make a list of all of the people on a piece of paper. You can give us nicknames or their relationship to you if you prefer (for example, 'my mother'). |  |
|  | [Loop questions SC02-SC09 for a random 10 people from SC01, or all if fewer than 10 reported] |  |
| sc02 | How many people are on your list? | integer |
| sc03 | How many of those people are members of your household? | integer |
| sc04 | What is your relationship to this person? | 0 "You live together" 1 "Neighbour" 2 "Relative" 3 "Work colleague" 4 "Friend" 5 "Stranger" 96 "Other" 98, "Prefer not to answer" |
| sc05 | How old do you think they are? | integer |
| sc06 | Are they male or female? | 1 "Male" 2 "Female" 99 "Don't know" 98 "Prefer not to answer" |
| sc07 | How much time did you spend with them in total? | time |
| sc08 | Did you touch them, for instance with a handshake, a hug or a kiss? | 0 "No" 1 "Yes" 99 "Don't know" 98 "Prefer not to say" |
| sc09 | How often do you typically see them? | 0 "6 or 7 days a week" 1 "2-5 days a week" 2 "Once a week" 3 "Once a week to once a month" 4 "Less than once a month" 5 "Never before" 98 "Prefer not to answer" |
| **Social contacts 2020** |  |  |
| sc01 | We will now ask you to remember who you have been in contact with yesterday, between 5am yesterday and 5am today. We are only interested in direct contacts, which are people who you met in person and with whom you exchanged at least a few words, or with whom you had physical contact (e.g. a handshake, embracing, kissing, contact sports). Note that if you only spoke to someone over the phone or internet, they should not be included.<br>Please write the nickname of each other person in your household. Note that this nickname is only needed to make it easier for you to complete the survey, so please pick a nickname that will help you identify each household member later in the questionnaire. Nicknames are not visible to anyone outside of this survey. |  |
|  | [Loop over list from SC01 for SC02] |  |
| sc02 | Which of these household members did you have direct contact with <u>in person</u>, between 5am yesterday and 5am today? | Yes, No |
| sc03 | And what other people did you have direct contact with in person, between 5am yesterday and 5am today? Please think about anyone else you had direct contact with. This could include friends, family, work colleagues, or people you spoke to in shops and so on. The order in which you give these names does not matter. However, it is easiest to give them in chronological order, e.g. when I woke up, I saw Busi and Thabo at breakfast. I then drove to my work, where I met with Thandiwe and Sfiso. On my way back home, I stopped at a petrol station, where I had a brief chat with the shop assistant. Etc.<br>Please write the nickname of each person you had direct contact with. Note that this nickname is only needed to make it easier for you to complete the survey, so please pick a nickname that will help you identify each contact later in the questionnaire. Nicknames are not visible to anyone outside of this survey. |  |
|  | [Loop over list from SC03 for SC04-SC11] |  |
| sc04 | What age is NAME? Please give an estimate if you are not sure. | 0-4, 5-9, 10-14, 70-74, 75-79, 80-84, 85+ |
| sc05 | What gender is NAME? | 0 Male; 1 Female; 96 Other; 98 Prefer not to answer; 99 Don't know |
| sc06 | What is NAME's relationship to you? | 0 They are a family member who is not in my household; 1 They are someone I work with; 2 They are someone I go to school, college or university with; 3 They are a friend; 96 Other; 98 Prefer not to answer |
| sc07 | Before the coronavirus epidemic started, how often did you usually have direct contact with NAME? A direct contact is when you meet with this person in person and when you exchange at least a few words, or when you have physical contact (e.g. handshake, embracing, kissing, contact sports). Please do not include times that you speak to them over the phone or internet. | 0 Every day or almost every day; 1 About once or twice a week; 2 Every 2-3 weeks; 3 About once per month; 4 Less often than once per month; 5 Never met them before; 98 Prefer not to answer |
| sc08 | When you had direct contact with NAME yesterday, did you have | 0 Physical contact (any sort of skin-to-skin contact such as e.g. hand shaking, embracing or kissing); 1 Non-physical contact (you did not touch the person); 98 Prefer not to answer |
| sc09 | Where did you have direct contact with NAME? [Do not read out list] | 0 "Own home" 1 "Other house on plot" 2 "House off plot" 3 "Church" 4 "Clinic" 5 "Community building" 6 "Creche" 7 "Gym" 8 "Library" 9 "Mall / Shops" 10 "in Bar / Spotini / Nightclub" 11 "Spaza" 12 "School" 13 "Salon/Barber" 14 "In a tent" 15 "Counselling Centre" 16 "Guest House" 17 "Office" 18 "Workshop" 19 "Restaurant" 96 "Other" 98 "Prefer not to say" |
| sc10 | Please estimate the total amount of time you spent with [NAME] in person yesterday | time |
| sc11 | Was the time you spent with [NAME] yesterday inside or outside? [Select all that apply] | 0 Inside; 1 Outside; 2 Both |

The questions listed here are the relevant sections of the 2019 Umoya Omuhle survey and the 2020 Covid Social Contacts survey, and not the complete questionnaires in each case. \* In 2020 the times were 5am and 5pm. † Questions only asked in 2019.

### Supplementary Material 2. Extended methods for statistical analysis

#### Weighting

To ensure both comparability across surveys and meaningfulness of results we weighted all our observed data to match the demographic surveillance area census population from which respondents were drawn. Additionally, for the UO data, when using contact characteristics, for respondents naming more than 10 contacts, we resampled from the 10 random contacts upon which more detailed information was provided, up to the total number of reported contacts. For the 2020 CSC data, we weighted respondents to account for stratified sampling and non-response by age, sex, comorbidity status, to match the Vuk’uzazi respondent sample. We then further weighted by age and sex to account for Vuk’uzazi non-response, to reach the census distribution. The UO data was weighted to the same age and sex distribution. As no respondents aged 15-17 were interviewed in the UO survey, respondents aged 18-29 were assumed to be representative of 15-29 year olds in the main analysis. Finally, we weighted responses by day of week, so that weekdays accounted for 5/7 of all observations, and weekends the remainder.

Age-mixing matrices were made symmetrical by setting the estimated rate of contact between each person in age group *i* and age group *j* equal to mean of the rates reported by respondents in each age group, using the age distribution of the census population to calculate the rates.

#### Management of incomplete data

Some respondents reported not knowing some or all of their contacts’ ages. We excluded from all the age-mixing matrices and R0 estimates any respondent who gave this response for all their close contacts in a round (n=2 in UO, n=10 in CSC R1, n=6 in CSC R2). People with missing data for all of their close contacts had a slightly lower mean number of close contacts (mean of 1.5 vs 7.3 in UO,mean of 3.5 vs 4.0 in CSC R1, mean of 3.2 vs 4.0 in CSC R2; all unweighted); this may reflect them being less engaged with the survey or a genuinely lower number of contacts. For respondents giving only some ages, we upweighted the known-aged contacts for the respondents to replace missing-age ones in the ‘best estimate’ age-mixing matrices, and excluded missing-age contacts from the resampling process when estimating confidence intervals for the age mixing matrices and when estimating the relative R0 reduction (while keeping the total number of contacts to sample for each respondent equal to the total number of contacts reported).

For the transport and location analysis, we excluded respondents reporting no location in the past 24-hours (including their own home) as implausible (n=1 in UO; n=1 R1, n=6 R2 in CSC). We used the same weights as for the close contact analyses.

#### Sensitivity analyses

For the analysis incorporating children: Since we ignored contacts aged under 15 in our primary analysis, because they were not eligible for either survey, we incorporated children into our analysis using the data on contacts with children reported by adults to calculate contact rates between children and adults. As we had no data on contact rates between children, we used data from a previous social contact study in South Africa to estimate these, making the assumption that the ratio of contact rates between 0-14 year old and contact rates between 15-29 year olds was the same (Johnstone-Robertson, Mark et al. 2011). In estimating R0, we also assumed that the probability of infection per contact was 44% lower for children than for adults (Viner, Mytton et al. 2020).

For the analysis excluding 15-17 year olds: As exact contact ages were not collected in CSC, we assumed that the proportion of all contacts aged 15-19 who were aged 18-19 was proportional to the proportion of the census population aged 15-19 who were aged 18-19.

## Supplementary Material 3. Sensitivity analyses

Estimated percent reductions in R0
|  | 2019 to 2020 R1 |  | 2019 to 2020 R2 |  | 2020 R1 to 2020 R2 |  |
| --- | --- | --- | --- | --- | --- | --- |
|  | Mean (95% PR) | p-value | Mean (95% PR) | p-value | Mean (95% PR) | p-value |
| Primary estimate | 41.7% (13.6, 59.1%) | 0.004 | 45.1% (24.2, 60.8%) | <0.001 | 2.3% (-53.0, 43.5%) | 0.4 |
| 1. Day of week weights | 42.1% (15.1, 59.1%) | 0.002 | 47.6% (28.9, 61.8%) | <0.001 | 6.4% (-43.6, 44.4%) | 0.4 |
| 2. Exclude respondents with missing contact ages | 49.5% (37.9, 59.6%) | <0.001 | 45.0% (22.0, 61.5%) | <0.001 | -10.2%, (-63.4%, 27.6%) | 0.5 |
| 3. Exclude short duration contacts | 43.9% (14.3, 62.2%) | 0.004 | 46.3% (24.3, 62.8%) | <0.001 | -0.015 (-62.1, 45.0%) | 0.5 |
| 4. Include children | 40.1% (18.4, 54.0%) | <0.001 | 41.4% (23.7, 55.3%) | <0.001 | 0.007 (-42.6, 34.5%) | 0.5 |
| 5. Exclude 15-17 year olds | 46.6% (34.0, 51.0%) | <0.001 | 37.8% (6.3, 59.2%) | 0.001 | -17.8% (-84.9, 27.1%) | 0.5 |
| 6. June-August only in 2019 data | 41.1% (12.8, 58.8%) | 0.005 | 44.5% (23.3, 60.7%) | <0.001 |  |  |
| 7. Urban/rural weights | 42.0% (14.2, 59.3%) | 0.004 | 45.4% (24.6, 61.0%) | <0.001 |  |  |

**1.**
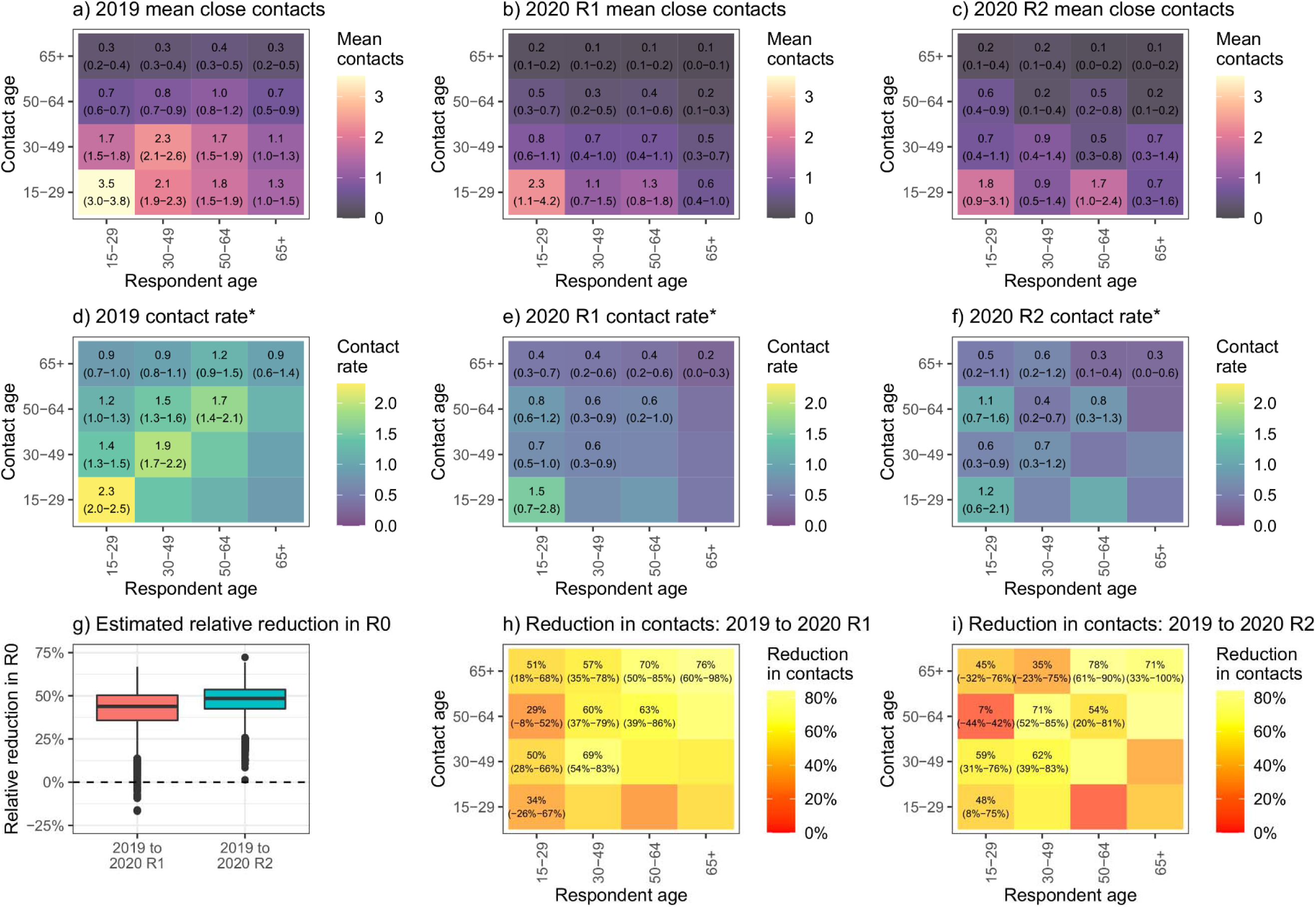
Day of week weights.

**2.**
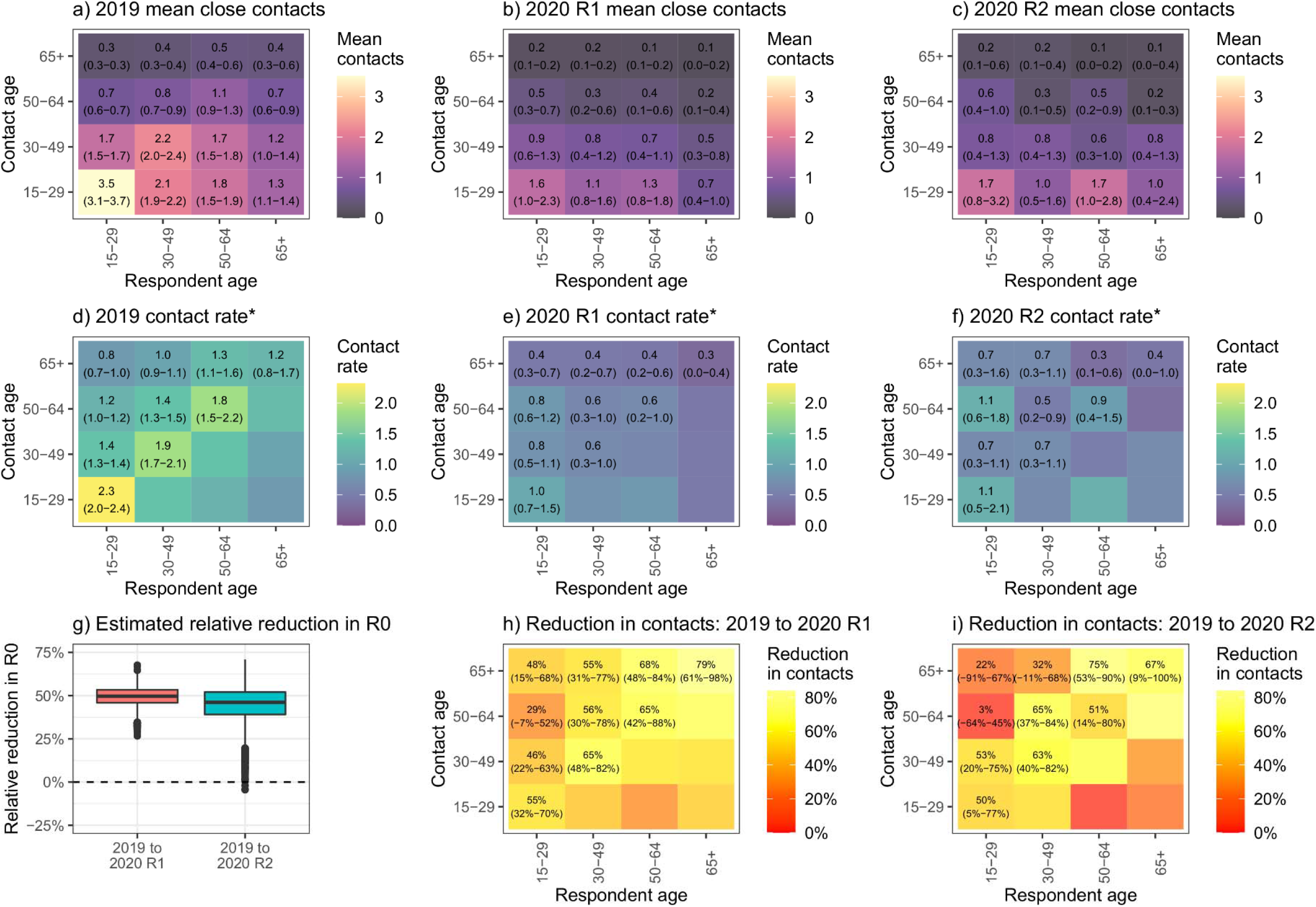
Exclude respondents with missing contact ages.

**3.**
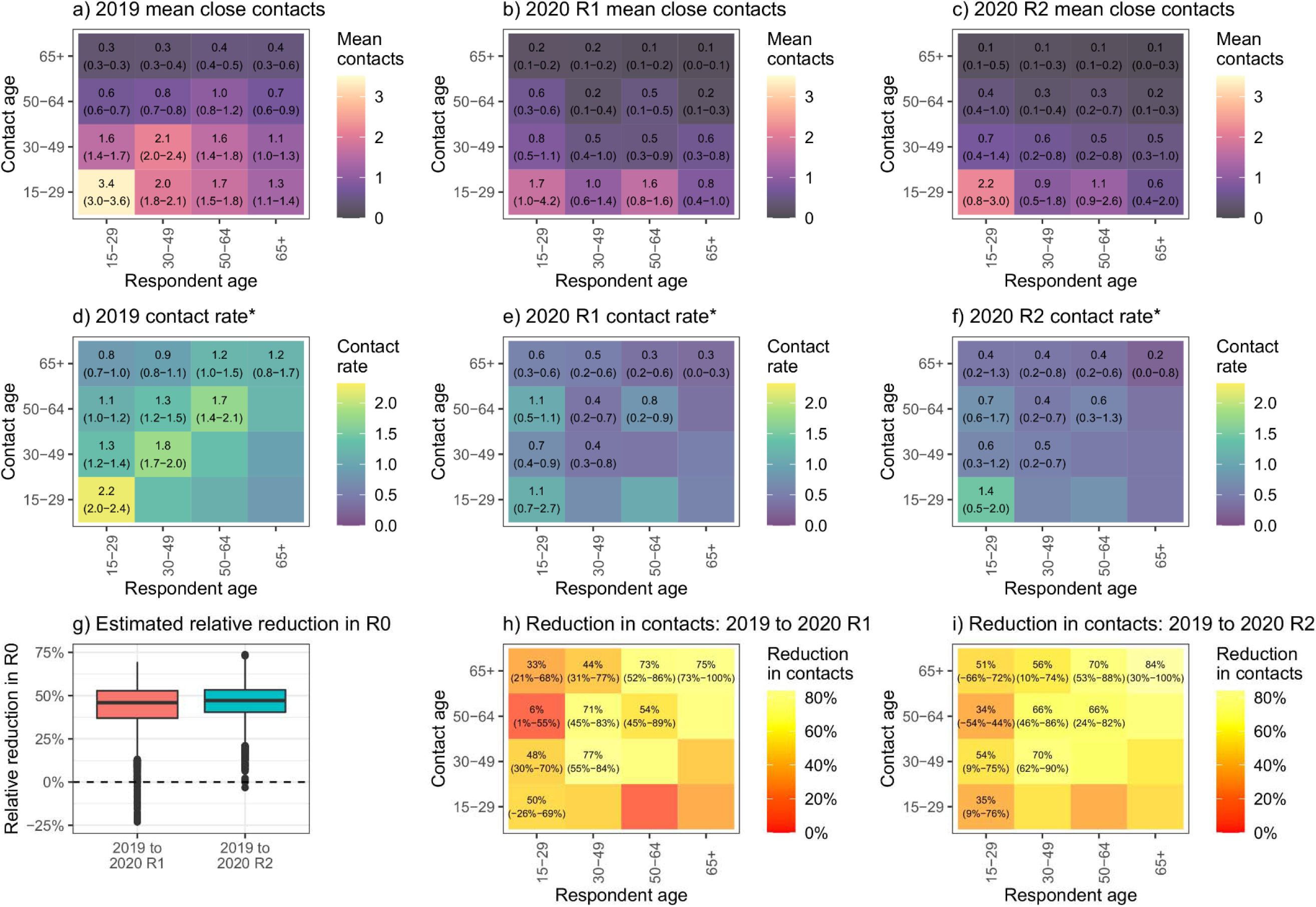
Exclude short duration contacts.

**4.**
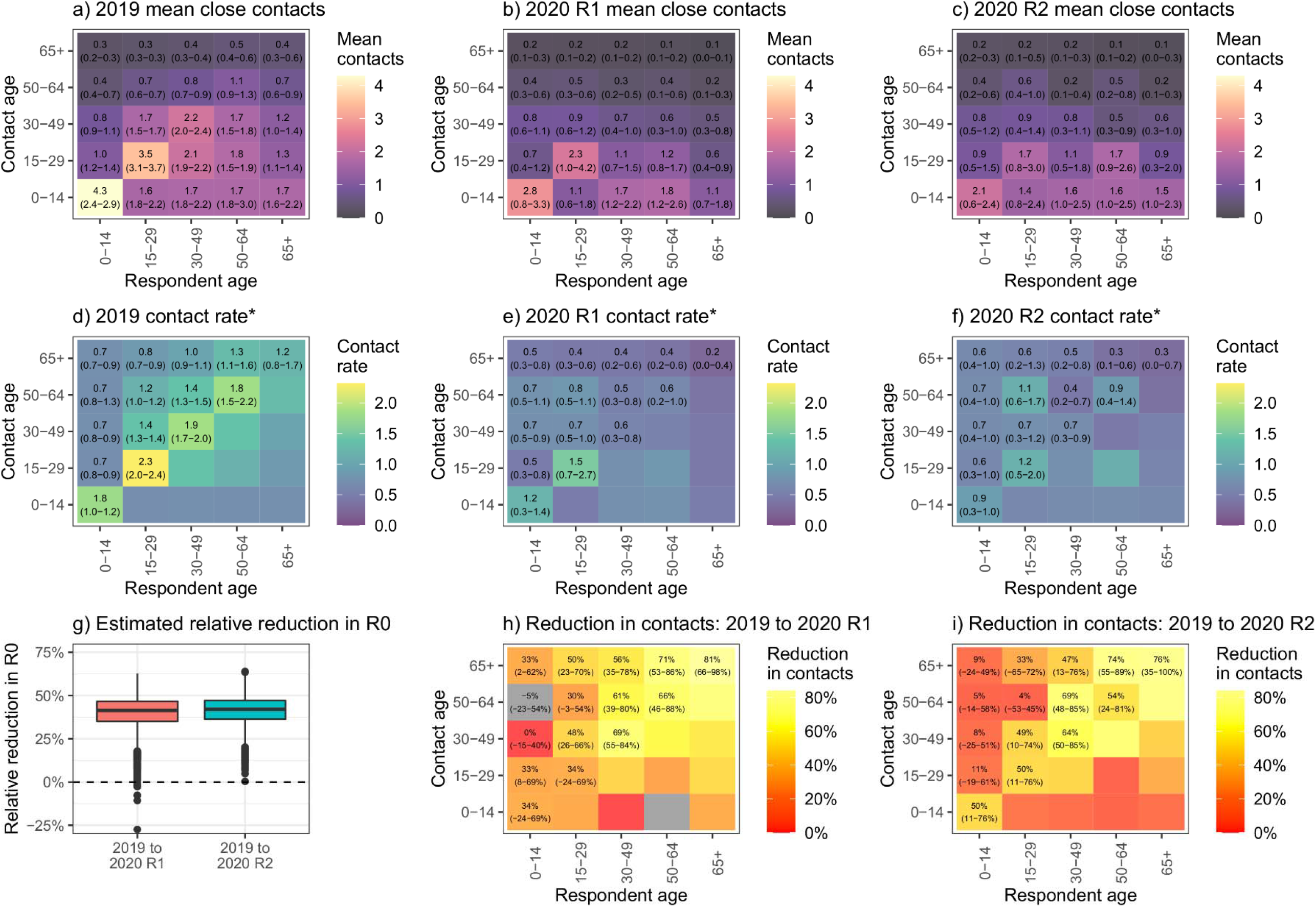
Include children. *The colour scales for a)-c) of this graph are different than those used in the other figures

**5.**
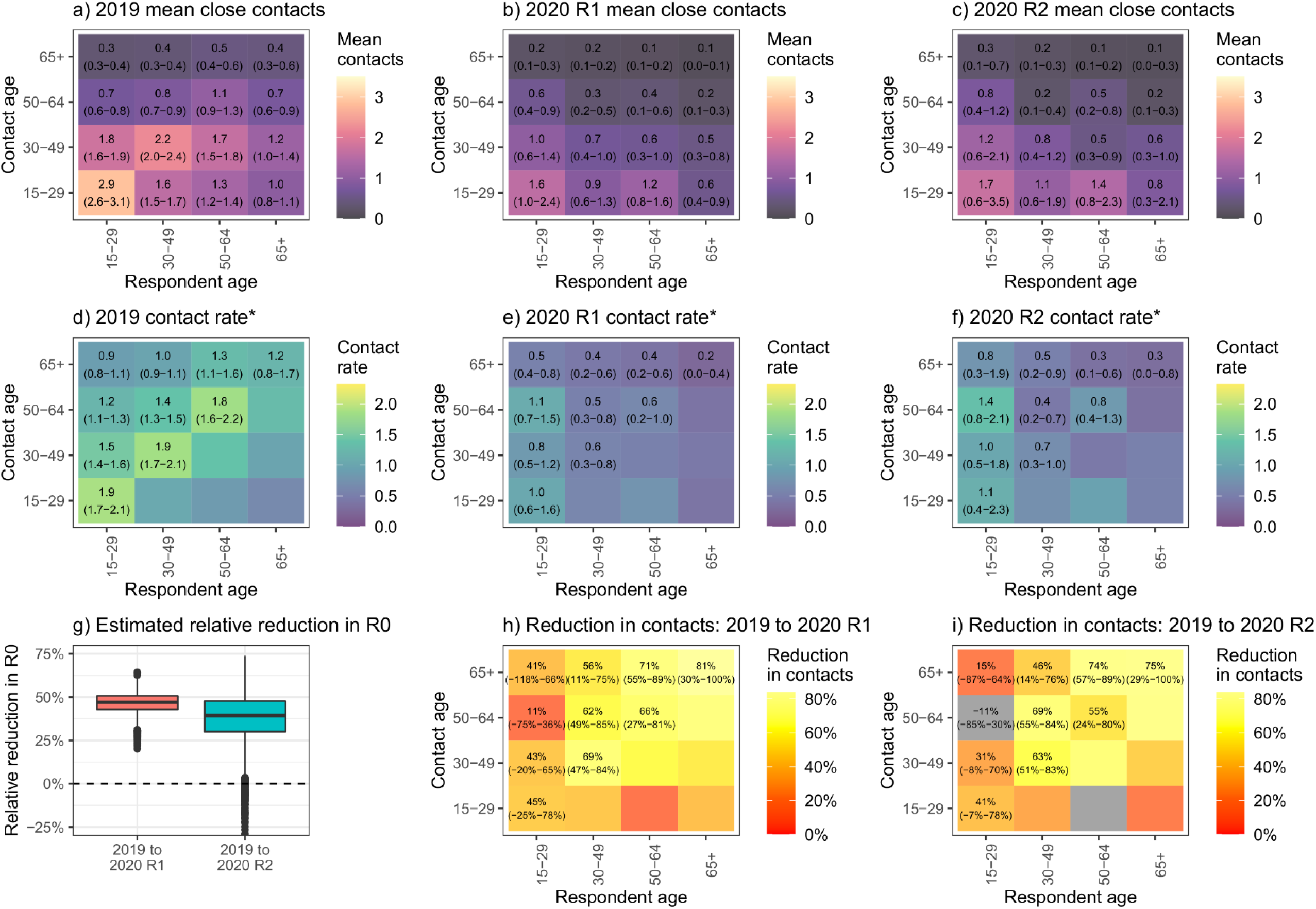
Exclude 15-17 year olds. * Grey in graph i) indicates an increase in contact numbers and rates

**6.**
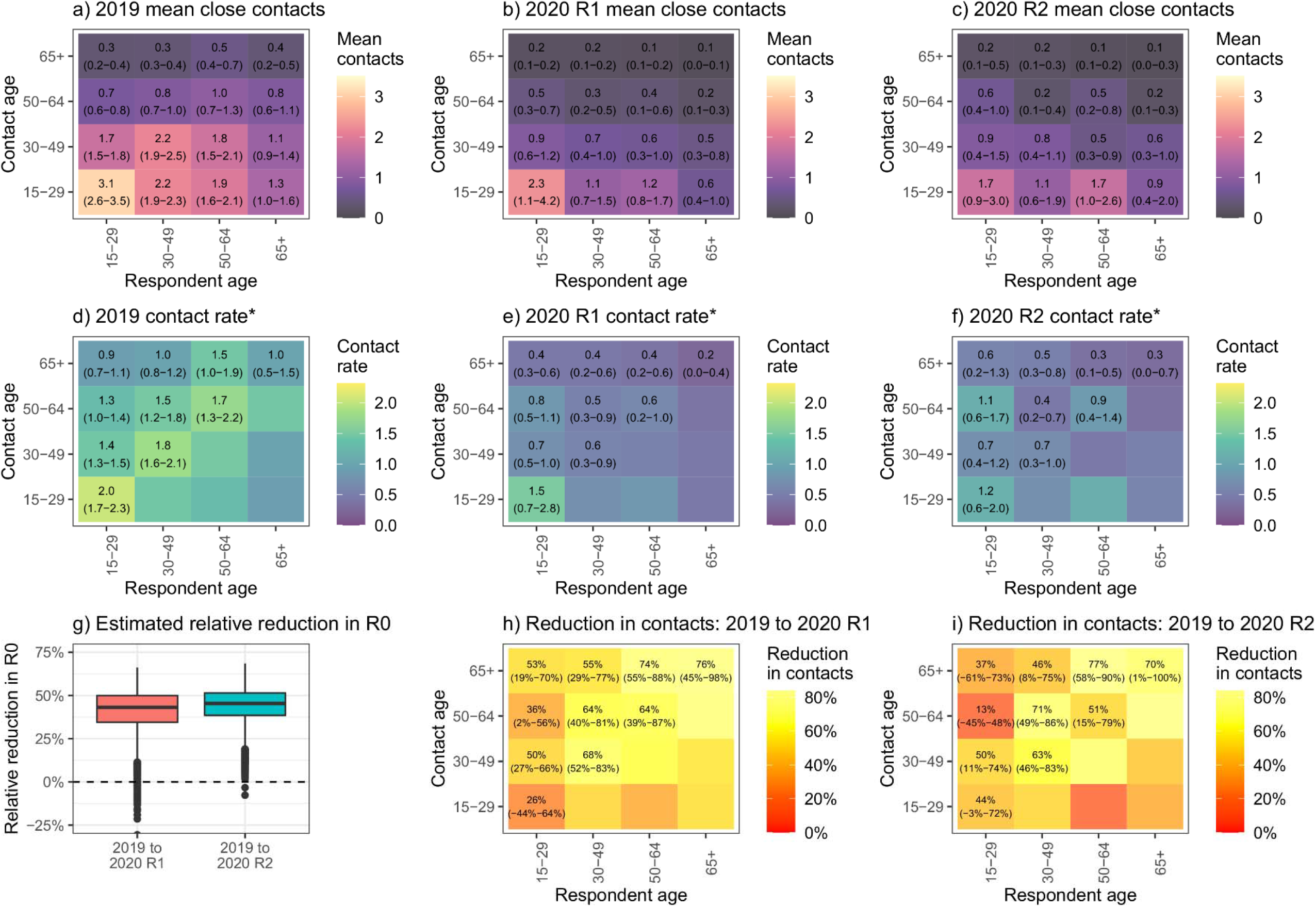
Only use UO data from June-August 2019.

**7.**
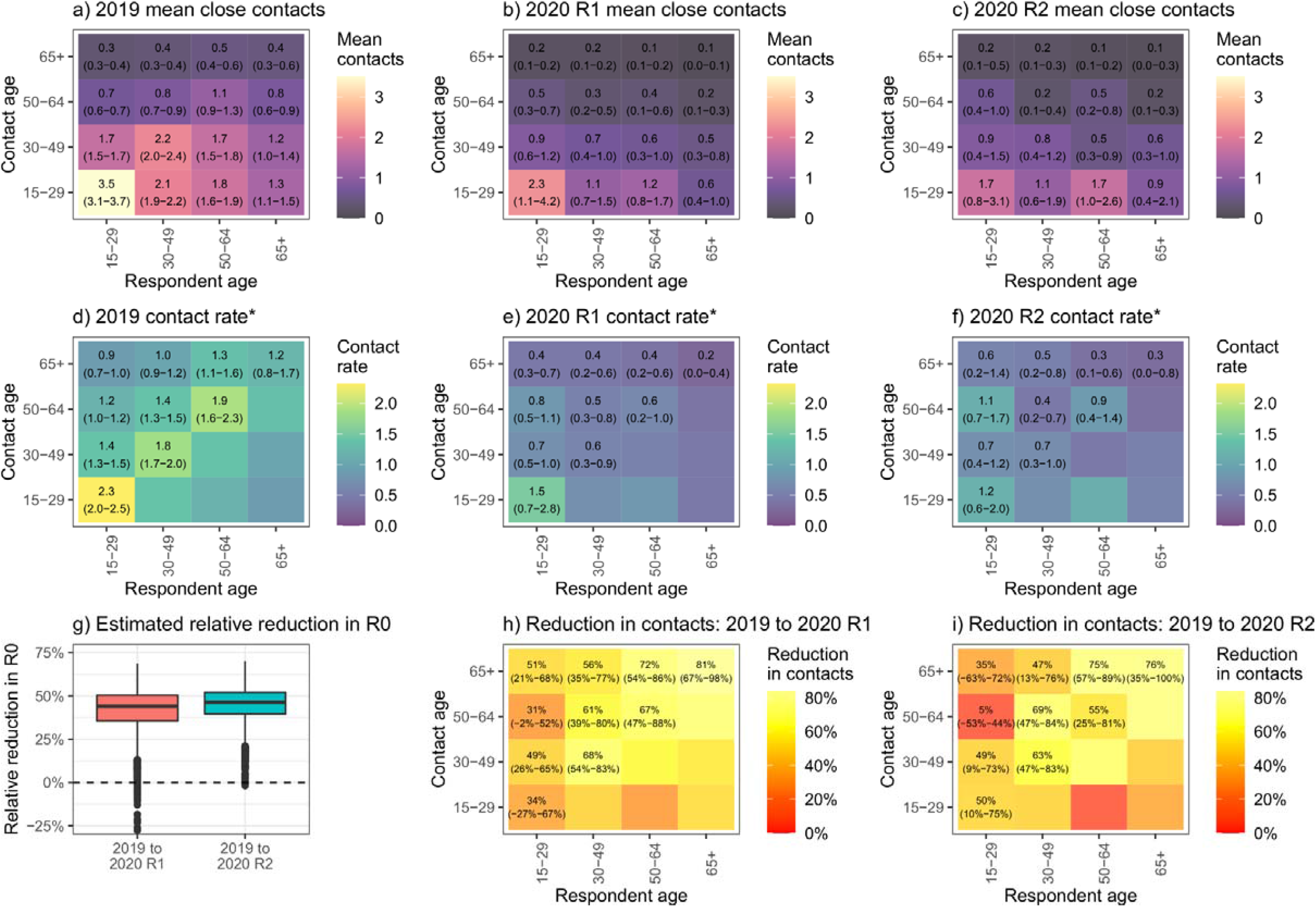
Weight the 2019 data to the proportion urban/peri-urban and rural in the census.

## Supplementary Material 4. Detailed description of ‘other locations’ from non-close contact questions

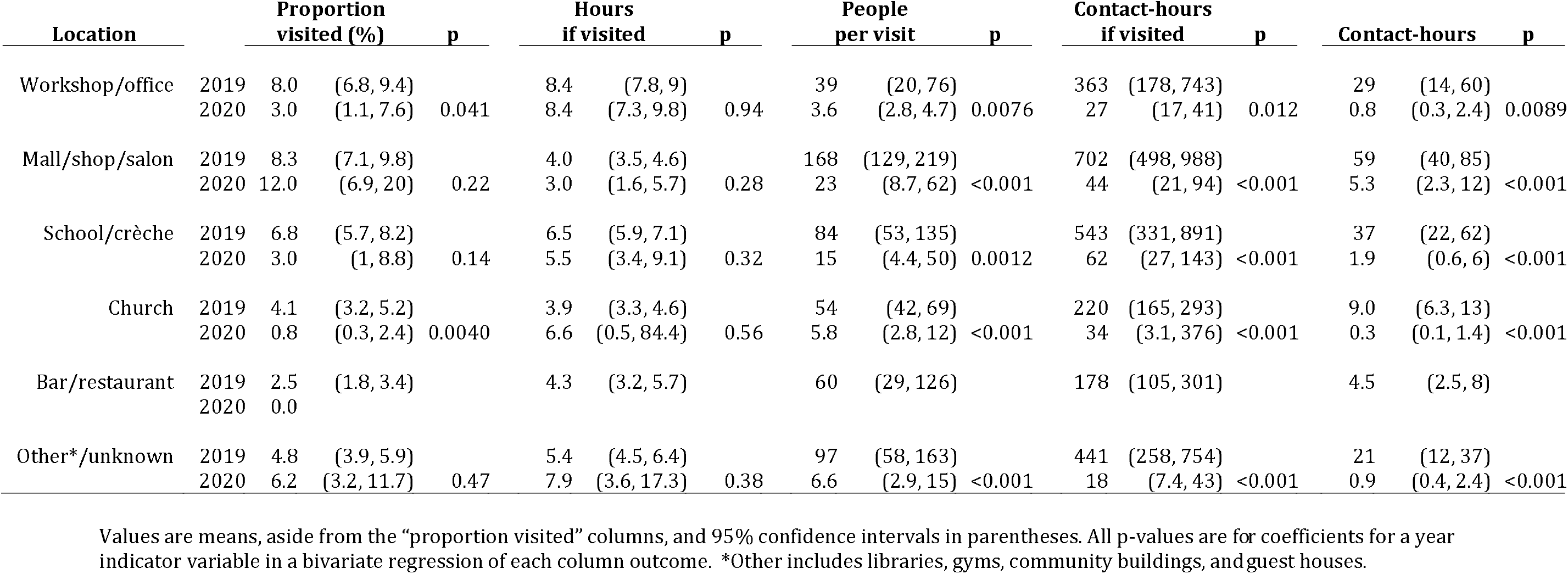

## Notes

### Competing Interest Statement

The authors have declared no competing interest.

### Author Declarations

Ethical approval for 2019 data collection was granted by the Biomedical Research Ethics Committee (REC) of the University of KwaZulu-Natal (UKZN) (BE662/17) and the London School of Hygiene & Tropical Medicine (14640); ethical approval for 2020 data collection was granted by UKZN BREC (BE290/16) and University College London REC (15231/013). Informed consent for participation was recorded in writing in 2019 and telephonically in 2020.

