## Supplementary material for "Impact of social distancing regulations and epidemic risk perception on social contact and SARS-CoV-2 transmission potential in rural South Africa: analysis of repeated cross-sectional surveys": STROBE checklist

STROBE Statement—checklist of items that should be included in reports of observational studies

| | Item No. | Recommendation | Page No. | Notes |
|---|---|---|---|---|
| **Title and abstract** | 1 | (*a*) Indicate the study's design with a commonly used term in the title or the abstract | 1 | In title |
| | | (*b*) Provide in the abstract an informative and balanced summary of what was done and what was found | 3 | |
| **Introduction** | | | | |
| Background/rationale | 2 | Explain the scientific background and rationale for the investigation being reported | 7-8 | |
| Objectives | 3 | State specific objectives, including any prespecified hypotheses | 8-9 | Last paragraph of Introduction |
| **Methods** | | | | |
| Study design | 4 | Present key elements of study design early in the paper | 9-10 | Second & third paragraphs of methods |
| Setting | 5 | Describe the setting, locations, and relevant dates, including periods of recruitment, exposure, follow-up, and data collection | 9 | First paragraph of methods |
| Participants | 6 | (*a*) *Cohort study*—Give the eligibility criteria, and the sources and methods of selection of participants. Describe methods of follow-up<br>*Case-control study*—Give the eligibility criteria, and the sources and methods of case ascertainment and control selection. Give the rationale for the choice of cases and controls<br>*Cross-sectional study*—Give the eligibility criteria, and the sources and methods of selection of participants | 9-10 | Second & third paragraphs of methods |
| | | (*b*) *Cohort study*—For matched studies, give matching criteria and number of exposed and unexposed<br>*Case-control study*—For matched studies, give matching criteria and the number of controls per case | NA | |
| Variables | 7 | Clearly define all outcomes, exposures, predictors, potential confounders, and effect modifiers. Give diagnostic criteria, if applicable | 10-12 | |
| Data sources/ measurement | 8* | For each variable of interest, give sources of data and details of methods of assessment (measurement). Describe comparability of assessment methods if there is more than one group | 10 | Fourth paragraph of methods |

| | | | | |
|---|---|---|---|---|
| Bias | 9 | Describe any efforts to address potential sources of bias | 11 & Supplementary Material 2 | We weighted all samples back to a recent population census, allowing for both sampling and non-response biases |
| Study size | 10 | Explain how the study size was arrived at | 9-10 | |
| Quantitative variables | 11 | Explain how quantitative variables were handled in the analyses. If applicable, describe which groupings were chosen and why | 11 | |
| Statistical methods | 12 | (*a*) Describe all statistical methods, including those used to control for confounding | 11-13 | |
| | | (*b*) Describe any methods used to examine subgroups and interactions | NA | |
| | | (*c*) Explain how missing data were addressed | NA | |
| | | (*d*) *Cohort study*—If applicable, explain how loss to follow-up was addressed<br>*Case-control study*—If applicable, explain how matching of cases and controls was addressed<br>*Cross-sectional study*—If applicable, describe analytical methods taking account of sampling strategy | 11 & Supplementary Material 2 | Inverse probability weighting to account for sampling back to study population |
| | | (*e*) Describe any sensitivity analyses | 12-13 | Last paragraph of methods |
| **Results** | | | | |
| Participants | 13* | (a) Report numbers of individuals at each stage of study—eg numbers potentially eligible, examined for eligibility, confirmed eligible, included in the study, completing follow-up, and analysed | 13 | First paragraph of results |
| | | (b) Give reasons for non-participation at each stage | 13 | First paragraph of results |
| | | (c) Consider use of a flow diagram | | |
| Descriptive data | 14* | (a) Give characteristics of study participants (eg demographic, clinical, social) and information on exposures and potential confounders | Table 1 | |
| | | (b) Indicate number of participants with missing data for each variable of interest | | |
| | | (c) *Cohort study*—Summarise follow-up time (eg, average and total amount) | | |
| Outcome data | 15* | *Cohort study*—Report numbers of outcome events or summary measures over time | | |
| | | *Case-control study*—Report numbers in each exposure category, or summary measures of exposure | | |
| | | *Cross-sectional study*—Report numbers of outcome events or summary measures | Table 2, Figure 1 | |

| | | | | |
|---|---|---|---|---|
| Main results | 16 | (*a*) Give unadjusted estimates and, if applicable, confounder-adjusted estimates and their precision (eg, 95% confidence interval). Make clear which confounders were adjusted for and why they were included | 13-14 | |
| | | (*b*) Report category boundaries when continuous variables were categorized | NA | |
| | | (*c*) If relevant, consider translating estimates of relative risk into absolute risk for a meaningful time period | NA | |
| Other analyses | 17 | Report other analyses done—eg analyses of subgroups and interactions, and sensitivity analyses | 15 | |
| **Discussion** | | | | |
| Key results | 18 | Summarise key results with reference to study objectives | 15 | First paragraph of discussion |
| Limitations | 19 | Discuss limitations of the study, taking into account sources of potential bias or imprecision. Discuss both direction and magnitude of any potential bias | 18 | |
| Interpretation | 20 | Give a cautious overall interpretation of results considering objectives, limitations, multiplicity of analyses, results from similar studies, and other relevant evidence | 16-17 | |
| Generalisability | 21 | Discuss the generalisability (external validity) of the study results | 17, 18 | |
| **Other information** | | | | |
| Funding | 22 | Give the source of funding and the role of the funders for the present study and, if applicable, for the original study on which the present article is based | 2 | |

*Give information separately for cases and controls in case-control studies and, if applicable, for exposed and unexposed groups in cohort and cross-sectional studies.
